# Analyses using multiple imputation need to consider missing data in auxiliary variables

**DOI:** 10.1101/2023.12.11.23299810

**Authors:** Paul Madley-Dowd, Elinor Curnow, Rachael A. Hughes, Rosie Cornish, Kate Tilling, Jon Heron

## Abstract

Auxiliary variables are used in multiple imputation (MI) to reduce bias and increase efficiency. These variables may often themselves be incomplete. We explored how missing data in auxiliary variables influenced estimates obtained from MI. We implemented a simulation study with three different missing data mechanisms for the outcome. We then examined the impact of increasing proportions of missing data and different missingness mechanisms for the auxiliary variable on bias of an unadjusted linear regression coefficient and the fraction of missing information. We illustrate our findings with an applied example in the Avon Longitudinal Study of Parents and Children. We found that where complete records analyses were biased, increasing proportions of missing data in auxiliary variables, under any missing data mechanism, reduced the ability of MI including the auxiliary variable to mitigate this bias. Where there was no bias in the complete records analysis, inclusion of a missing not at random auxiliary variable in MI introduced bias of potentially important magnitude (up to 17% of the effect size in our simulation). Careful consideration of the quantity and nature of missing data in auxiliary variables needs to be made when selecting them for use in MI models.

## Background

Missing data can lead to bias in effect estimates and reduced statistical efficiency. Types of missing data include missing completely at random (MCAR) where the probability of missing data does not depend on observed or unobserved values of analysis model variables, or any related variables, missing at random (MAR) where conditional on observed data, the probability of missing data is independent of unobserved data, and missing not at random (MNAR), where the probability of missing data is dependent on unobserved data even after conditioning on observed data ^1^.

Common strategies for handling missing data include complete records analysis (CRA) and multiple imputation (MI). CRA analyses only the subset with complete data for all variables in the analysis model. MI, on the other hand, creates multiple datasets with imputed values from predictive models (the imputation model), fits the analysis model in each imputed dataset, and combines effect estimates using Rubin’s rules ^2^. Under standard implementation, MI assumes each variable to be imputed is MAR, and that all imputation models are correctly specified and compatible with the analysis model ^3^ (e.g., the imputation models includes all variables in the analysis model including interactions and non-linear terms). Under a MAR mechanism MI can reduce bias and losses in efficiency from missing data. Unfortunately, testing whether data are MAR is not possible ^4^.

In MI, auxiliary variables are included in the imputation model but not the analysis model. They can improve the plausibility of the MAR assumption (if they predict the probability of missing data and predicting missing values, thereby reducing bias) or improve the precision of effect estimates (if they predict the missing values only) ^5–8^. For example, in a study assessing the effect of maternal smoking during pregnancy (the exposure) on offspring intelligence quotient (IQ) scores (the outcome), adjusted for confounders, IQ scores may be more likely to be missing for individuals with learning difficulties, leading to biased CRA estimates as the probability of missing data is dependent on the outcome ^9^, and biased MI estimate as the data are MNAR. Including an auxiliary variable such as linked educational records in the imputation model would improve the plausibility of the MAR assumption and reduce bias in MI estimates^10^.

Historically, it was recommended to use as many auxiliary variables in the imputation model as possible to “reduce the chance of omitting an important cause of missingness”, reduce bias and improve efficiency with minimal cost ^7^. This view has been challenged in its extreme by work showing that model performance degraded (in terms of bias and precision) as the number of included auxiliary variables approached the number of records with complete data ^8^. The inclusion of auxiliary variables can exacerbate or induce bias, for example, where the auxiliary variable is a collider and conditioning upon it would induce a dependency between the outcome and the probability of missing data ^11,12^. Consideration of the causal relationships between each variable in the analysis question and the missingness mechanism is therefore essential when deciding which auxiliary variables should be included in imputation models.

Potential auxiliary variables may be incomplete. The amount of incomplete auxiliary data will not affect a CRA, because auxiliary variables are not in the analysis model. However, completeness of auxiliary variables could affect 1) how well the MAR assumption has been met for incomplete variables in the analysis model, and 2) the quantity of random noise introduced by imputing missing values of auxiliary variables. Prior work has briefly explored the impact of missing data in an auxiliary variable but only investigated a single proportion of missing data (20%) and did not explore the impact of different missing data mechanisms^10^. Further research is needed to investigate the impact of both (i) increasing proportions of missing auxiliary data and (ii) different missing data mechanisms for auxiliary variables on bias and efficiency of estimates obtained from MI.

## Methods

As an applied example, we investigated the relationship between maternal smoking during pregnancy and offspring IQ at age 15 years in the Avon Longitudinal Study of Parents and Children (ALSPAC). The outcome (offspring IQ) was incomplete, and two auxiliary variables were available which are not in the main analysis model but are correlated with the outcome. These were IQ at age 8 and a continuous attainment score for secondary education. The former contained more missing data but had a higher correlation with the outcome than the latter. We wished to improve our decision making about which auxiliary variables to include in the imputation models when the auxiliary variables contain missing data.

To aid our understanding, we undertook a simulation study to explore the impact of increasing proportions of missing data in auxiliary variables under different missing data mechanisms. All analyses were conducted using Stata version 17.

### Applied example

Data were taken from ALSPAC ^13–15^ which recruited 14,541 pregnant women resident in Avon, United Kingdom with expected dates of delivery 1st April 1991 to 31st December 1992. Of these pregnancies, 13,988 children were alive at 1 year of age. Inclusion criteria were being from a singleton pregnancy and surviving to one year of age, leaving a sample size of 13,826. To simplify this example, we excluded participants if they had missing data for any of the confounders to isolate the influence of auxiliary variables for the outcome. Participants with missing data in auxiliary variables were retained. Following exclusions our sample size was 11,780.

The substantive analysis was a linear regression of offspring IQ at age 15 on maternal smoking in pregnancy adjusted for the confounders maternal age, education, parity, and offspring sex. Maternal smoking during pregnancy was captured as a self-reported binary variable at 18 weeks’ gestation and offspring IQ was measured using the Wechsler Abbreviated Scale of Intelligence at age 15 years^16^. The auxiliary variables used in the imputation models were IQ at age 8, measured using the Wechsler Intelligence Scale for Children - III ^17^ and the capped Key Stage 4 (KS4) point score, a measure of educational attainment at age 16 obtained from linkage to the National Pupil Database and equal to the total score of an individual’s top eight GCSE or equivalent qualifications ranked in terms of points. A directed acyclic graph (DAG) ^18^ of the assumed relationships between variables and indicators for missing data is presented in the supplement along with justification for these assumptions.

We performed fully conditional specification (FCS) MI ^19^ using Stata’s *mi impute chained* command with 1000 imputations and 25 burn-in iterations. Six imputation models were investigated: i) excluding all auxiliary variables, ii) including IQ at age 8, iii) including KS4 score, iv) including IQ at age 8 and KS4 score, v) including KS4 score cubed and a multiplicative term for KS4 score cubed and maternal education and vi) including both IQ at age 8 and KS4 score cubed with a multiplicative term between KS4 score cubed and maternal education. In model v and vi KS4 score was imputed using the cube roots of IQ and a multiplicative term with maternal education. Models v and vi were included to highlight the importance of correctly specifying the imputation model as previous work has shown that IQ and KS4 attainment were related non-linearly and the relationship varied according to maternal education ^20^.

We report the effect estimate and standard error of the exposure coefficient (i.e., the effect of maternal smoking during pregnancy on offspring IQ at age 15) and the fraction of missing information (FMI). The FMI is a parameter-specific measure that quantifies the loss of information due to missing data while accounting for information recovered by MI ^2,21^. Values of FMI range between 0 and 1. Values close to 1 indicate high variability between imputed data sets, meaning that observed data in the imputation model does not provide much information about the missing data. A large number of imputations was used as the estimate of the FMI is highly variable (see the supplement of ^22^).

### Simulation study

#### Data generation model

Table 1 presents a summary of all simulation design factors and levels investigated. We generated 1,000 independent simulated datasets of sample size 1,000. Each dataset consisted of continuous variables Y (the outcome), X (the exposure) and Z (an auxiliary variable correlated with Y but not X), simulated from a multivariate normal distribution. Each variable had nonzero mean ^(̅^Y=6, ^̄^X=-3, ^Z̅^=2) and variance 1. The correlation of Y and X was held constant at 0.6 while the correlation of Z and Y was varied between 0.1 and 0.7 in increments of 0.2.

**Table 1:**
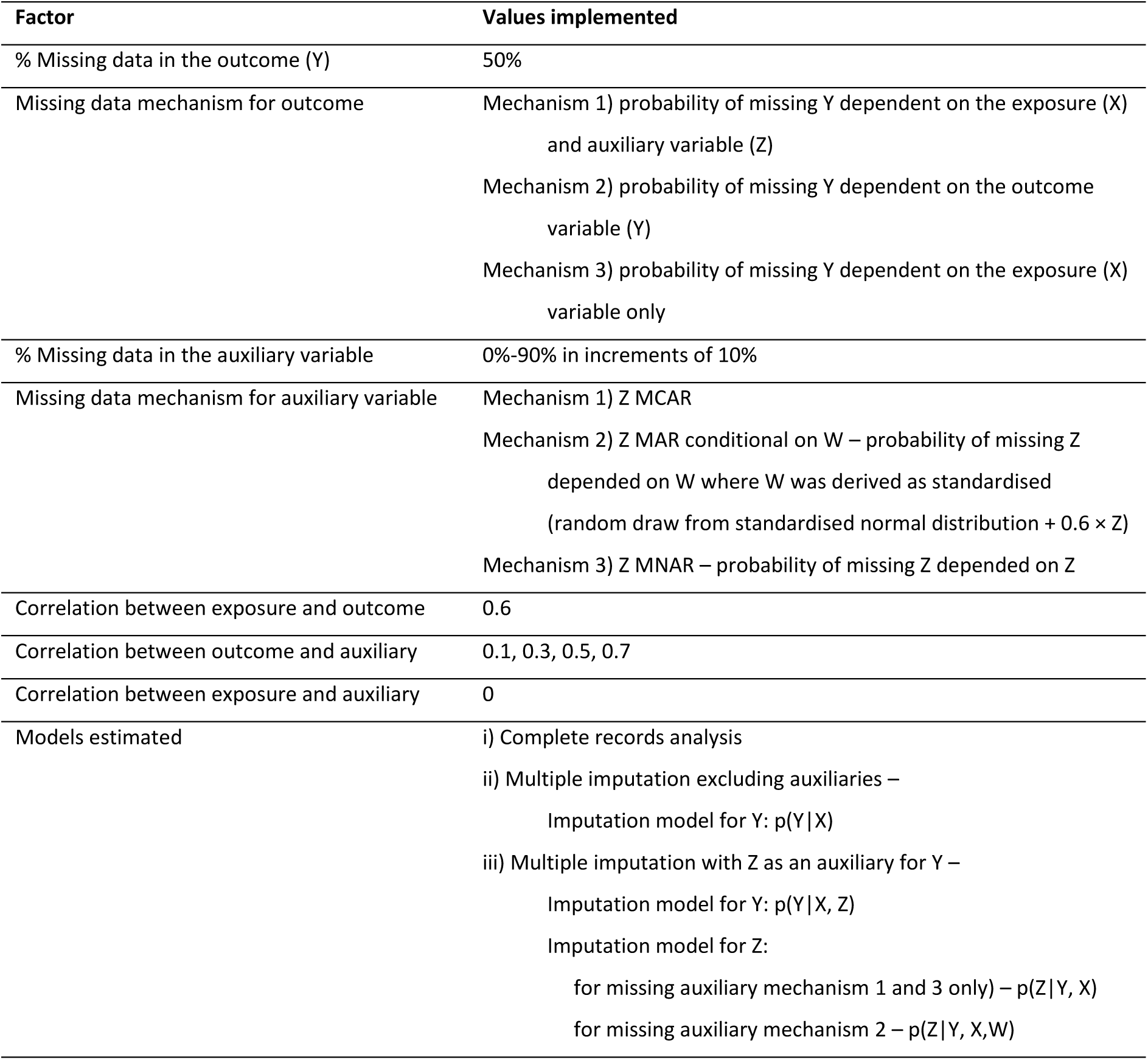
Simulation design factors and chosen levels. The simulation study was conducted under every combination of values.

We simulated 50% missing data in the outcome under three different mechanisms, displayed in Figure 1 as DAGs. The missing outcome mechanisms were as follows:

1. Y MAR conditional on complete X and Z
2. Y MNAR, missing dependent on its own value
3. Y MAR conditional on complete X

**Figure 1:**
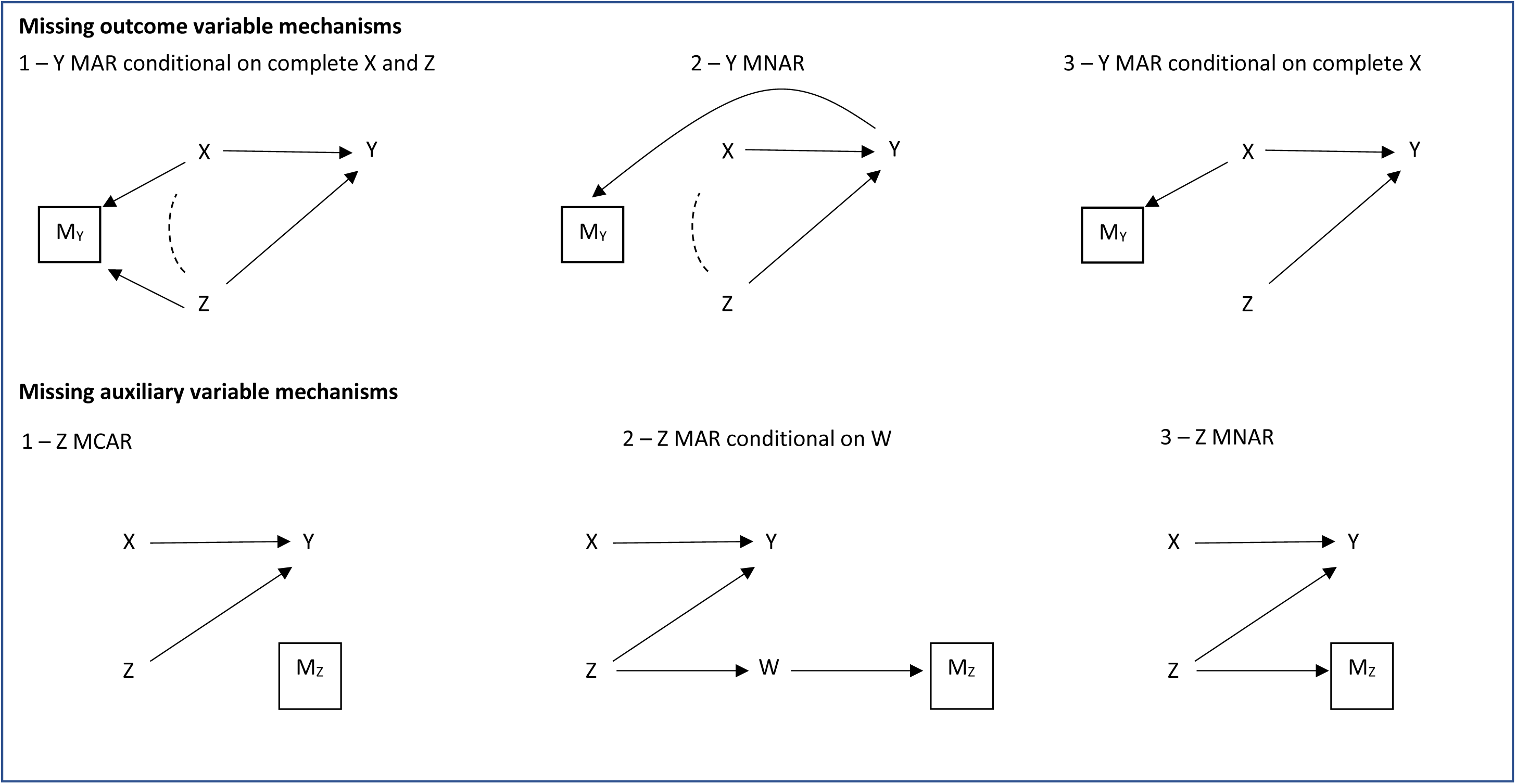
Directed acyclic graphs of the missing data mechanisms used for the outcome Y and auxiliary variable Z. Dashed arrows represent induced correlations that do not exist in the complete data. In outcome missingness mechanism 1, complete records analysis of a regression of Y on X is biased due to conditioning on the collider M_Y_ which induces correlation between X and Z. In outcome missingness mechanism 2, bias in complete records analysis occurs in the same way following conditioning on a child variable of the outcome Y which is a collider for X and Z. In outcome missingness mechanism 3, complete records analysis is unbiased. The missing data mechanisms for the outcome and auxiliary variables are presented separately for simplicity but can be combined to reflect the missing data in both variables.

In missing outcome mechanism 1 Y was set to missing if the cumulative distribution function (CDF) of X or Z was less than √0.5. In missing outcome mechanism 2 Y was set to missing if the CDF of Y was less than 0.5. This mechanism was chosen to reflect the scenario where data in IQ are likely to be missing dependent on intelligence, and we are using a proxy outcome such as educational attainment score to make the MAR assumption more plausible. In missing outcome mechanism 3 Y was set to missing if the CDF of X was less than 0.5. In outcome mechanisms 1 and 2, CRA is biased and MI using Z as an auxiliary would be implemented to reduce bias and improve efficiency. For outcome mechanism 3, CRA is unbiased, and MI using Z would be implemented for the purposes of improving efficiency only.

Missing data in the auxiliary variable was varied between 0 and 90%, simulated using 3 different missing data mechanisms (see Figure 1):

1. Z MCAR
2. Z MAR conditional on a completely observed variable W (correlated with Z)
3. Z MNAR, missing dependent on its own value

In missing auxiliary mechanism 1 Z was set to missing if a random draw from the uniform distribution bounded by 0 and 1 was less than μ, where μ was varied between 0 and 0.9 in increments of 0.1. In missing auxiliary mechanism 2 W was equal to 0.6 times Z plus a draw from a standard normal distribution (and then standardised) and Z was set to missing if the CDF of W was less than μ. In missing auxiliary mechanism 3 Z was set to missing if the CDF of Z was less than μ.

#### Analysis and imputation models

The analysis model consisted of a linear regression of Y on X. The true value of the effect estimate for X was equal to 0.6. For each scenario, the effect of X on Y was estimated using i) CRA and ii) MI where Z was not included as an auxiliary variable – missing Y imputed under the conditional model p(Y|X). We further estimated the effect of X on Y using iii) MI analysis where Z was included as an auxiliary variable for Y – missing Y imputed under the conditional model p(Y|X,Z). For missing auxiliary mechanism 1 and 3, Z was imputed using the conditional model p(Z|Y,X). For missing auxiliary mechanism 2, where W acted as a proxy for Z (but not Y), Z was imputed using the conditional model p(Z|Y, X, W). FCS MI was performed using Stata’s *mi impute chained* command using 100 imputations and 10 burn-in iterations.

#### Simulations and metrics

We report the bias and FMI (for MI models) of the effect estimate of X on Y for each scenario. Bias was calculated using the *simsum* command in Stata ^23^ and FMI for the exposure coefficient was calculated using Stata’s *mi estimate* command.

#### Sensitivity analyses

We conducted several sensitivity analyses exploring the impact of i) the proportion of missing data in the outcome and ii) the strength of association between X and Y relative to Z and Y. We further explored the impact of omitting W from the imputation model under missing auxiliary mechanism 2. These analyses and their results are detailed in the supplement.

## Results

### Applied example

Supplementary Table S1. shows descriptive statistics for the sample. Data in IQ at age 15 (outcome) were missing for 60% of the sample, in IQ at age 8 (auxiliary) for 44.2% and in KS4 scores (auxiliary) for 25.1%. The observed correlation between IQ at age 8 and 15 was 0.63, and between KS4 score and IQ age 15 was 0.59. Maternal smokers during pregnancy were more likely to have higher parity, lower levels of education and slightly more likely to have male offspring.

Table 2 shows results for our applied example. We discuss the effect estimates here and the FMI in the supplement. In CRA, smoking during pregnancy was associated with a 0.87-point reduction (95%CI=-1.82 to +0.08) in offspring IQ score at age 15. Comparable effect estimates were found between CRA and MI excluding auxiliary variables (MI model i). Using IQ at age 8 as an auxiliary in MI model ii resulted in a greater effect size of a 1.13-point reduction (95%CI=-2.03 to −0.23). Use of KS4 score as an auxiliary (model iii) further increased the effect size to a 1.99-point reduction (95%CI=- 2.87 to −1.10). Using both KS4 score and IQ at age 8 (model iv) provided a similar effect size and standard error to use of KS4 only. MI with KS4 score including non-linear terms (model v) attenuated the effect estimate to a 1.40-point reduction (95%CI=-2.29 to −0.52). Including IQ at age 8 and KS4 score with non-linear terms (model vi) resulted in a slightly higher estimate of a 1.49-point reduction (95%CI= −2.31 to −0.68) with the lowest standard error of all models.

**Table 2:** Model results for applied MI analyses.

| Model | Exposure coefficient (s.e.) <sup>a</sup> | 95% CI | FMI for exposure coefficient | Largest FMI of all covariates |
| --- | --- | --- | --- | --- |
| CRA | -0.87 (0.485) | -1.82, 0.08 | - | - |
| i) MI excluding auxiliaries | -0.86 (0.485) | -1.81, 0.09 | 0.70 | 0.70 |
| ii) MI with IQ at age 8 as an auxiliary | -1.13 (0.459) | -2.03,-0.23 | 0.66 | 0.66 |
| iii) MI with KS4 score as an auxiliary | -1.99 (0.453) | -2.87,-1.10 | 0.61 | 0.61 |
| iv) MI with IQ at age 8 and KS4 score as auxiliaries | -1.93 (0.444) | -2.80,-1.06 | 0.61 | 0.61 |
| v) MI with KS4 score cubed as an auxiliary (including multiplicative term <sup>b</sup>) | -1.40 (0.452) | -2.29,-0.52 | 0.64 | 0.64 |
| vi) MI with IQ at age 8 and KS4 score cubed as auxiliaries (including multiplicative term <sup>b</sup>) | -1.49 (0.414) | -2.31,-0.68 | 0.58 | 0.58 |

<sup>a</sup> Mean difference in IQ at age 15 among offspring of maternal smokers during pregnancy compared to non-smokers during pregnancy
<sup>b</sup> Multiplicative term between KS4 score cubed and maternal education

The true effect of maternal smoking in pregnancy on offspring IQ is unknown, as are the quantity of bias in CRA and which MI model achieved the greatest reduction in bias. Based on our DAG of the assumed relationship between variables (see supplement), CRA is likely biased due to the relationship between IQ and the probability of missing data in IQ ^9^. We were unable to completely mitigate this but used auxiliaries to proxy for missing IQ score and reduce bias ^10,20^. We consider model vi to provide the best effect estimate as it a) captures the non-linear relationship between the auxiliary KS4 score and the outcome and b) contains two auxiliary variables that are missing for different groups of people (see supplement for further detail) meaning that proxy information is available for a wider coverage of the distribution of missing outcome values.

### Simulation study

As expected, CRA was biased under missing outcome mechanisms 1 and 2 and unbiased under mechanism 3. As the correlation between Y and Z increased, the bias in CRA increased for outcome mechanism 1 but remained constant for outcome mechanism 2 (see Supplementary Table S2). Models excluding auxiliaries were biased to a similar extent as CRA under mechanism 1 and 2 and both were unbiased under mechanism 3.

Results for the FMI are presented in the supplement. We display plots of bias for all outcome and auxiliary missing data mechanism combinations in Figure 2. Bias is presented as relative to CRA for outcome missing data mechanisms 1 and 2 (as CRA was biased) and absolute bias for outcome mechanism 3 (as CRA was unbiased).

**Figure 2:**
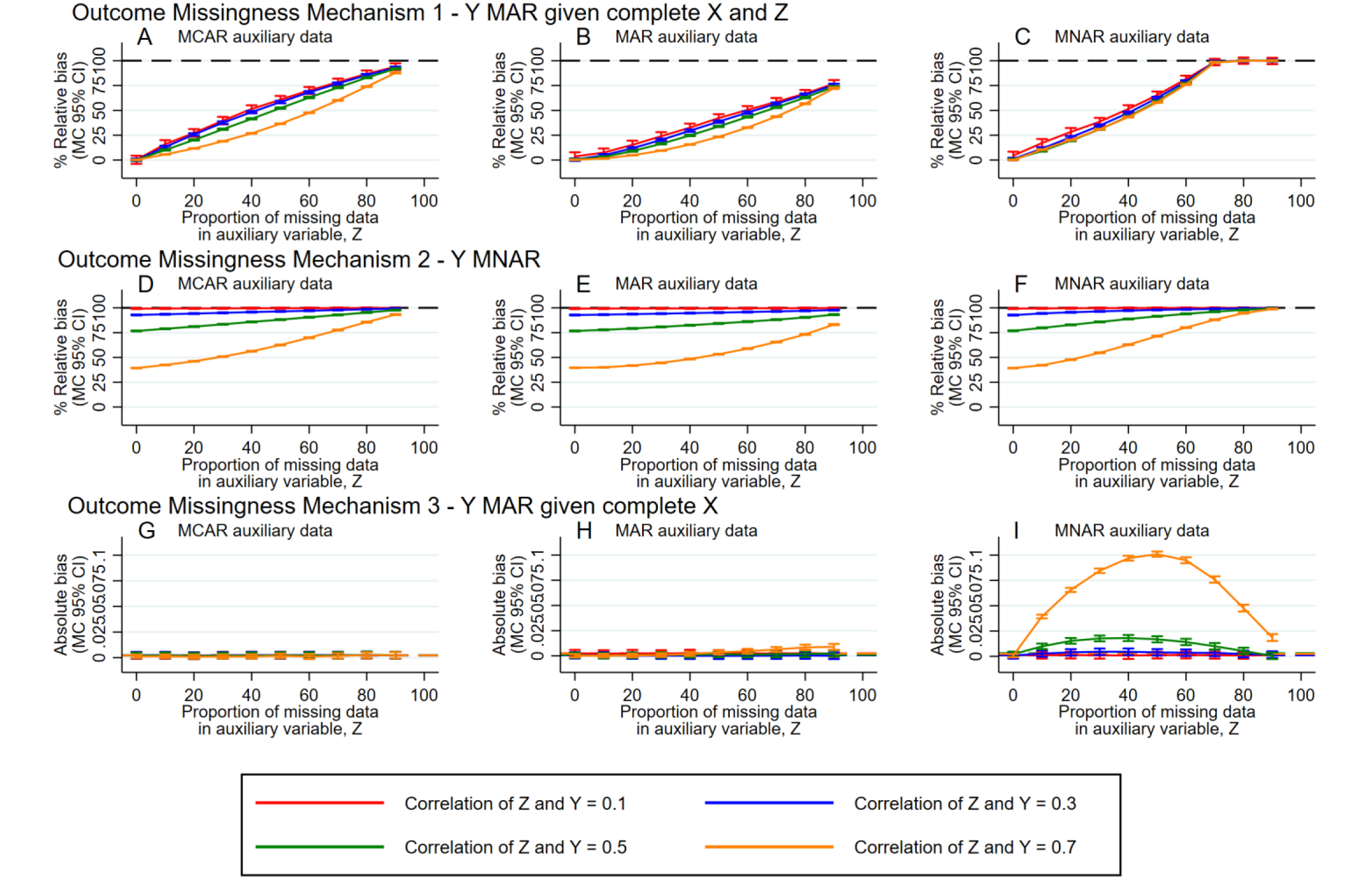
Plots of bias of the effect estimate of the exposure, X, across simulations against the proportion of missing data in the auxiliary variable, Z, for each level of correlation between the auxiliary, and outcome, Y. Bias is presented as relative to CRA for outcome missingness mechanisms 1 and 2, where CRA is expected to be biased, and absolute bias for outcome missingness mechanism 3, where CRA is expected to be unbiased. Solid lines correspond to bias in MI analysis including auxiliary variables while dashed lines correspond to bias in CRA which was approximately equivalent to MI excluding auxiliary variables. CI = confidence interval, MC = Monte Carlo, MCAR = missing completely at random, MAR = missing at random, MNAR = missing not at random.

For missing outcome mechanism 1 (MAR outcome given complete exposure and auxiliary; Figure 2 A-C), where there was no missing data in the auxiliary variable, MI models including auxiliaries resulted in no bias under all auxiliary missing data mechanisms. As the proportion of missing auxiliary data increased, bias in MI models approached that of CRA. For a given proportion of missing auxiliary data, bias was closer to that of CRA for auxiliary variables with weaker correlation with the outcome than those with stronger correlation. Bias relative to CRA was greater for MCAR auxiliary data (Plot A) than MAR auxiliary data (Plot B) at each proportion of missing data, with bias for MAR auxiliary data remaining below 80% of CRA even at 90% missing data in the auxiliary. For MNAR auxiliary data (Plot C), bias reached that of CRA by 70% missing auxiliary data.

Under missing outcome mechanism 2 (MNAR outcome where the auxiliary acts as a proxy; Figure 2 D-F) MI including auxiliary variables never completely removed bias. Bias reduction was greater when the auxiliary variable strongly predicted the missing outcome – the weakest correlation (0.1) did not result in any bias reduction. Bias approached that of CRA as the missing data in the auxiliary variable increased. The missing data mechanism for the auxiliary variable did not have a large influence on the magnitude of bias reduction.

Under missing outcome mechanism 3 (MAR outcome given complete exposure; Figure 2 G-I) no bias was observed when auxiliary data was MCAR (Plot G). A small quantity of bias was found for MAR auxiliary data (Plot H) at large proportions of missing data when the outcome and auxiliary were strongly correlated. When the auxiliary variable was MNAR (Plot I) bias was observed in MI models including auxiliary variables. Bias was larger for auxiliaries with stronger correlation with the outcome and increased as the proportion of missing auxiliary data increased from 0% to 50% but then declined as the proportion rose from 50% to 90%. The absolute bias reached a maximum of around 0.1, or 17% of the true effect size.

## Discussion

In this simulation study we demonstrated that increasing proportions of missing data in auxiliary variables limits their ability to reduce bias and recover information lost to missing data (as measured by the FMI) when included in imputation models. When MI is used for the purposes of improving efficiency only (i.e., there is no bias in CRA), including a MNAR auxiliary variable in an imputation model can induce bias. This bias was greatest when using an auxiliary that was a strong predictor of the outcome. Guidance on MI advises that such an auxiliary variable is desirable^6^. We do not challenge this guidance, but we highlight the importance of considering the quantity and causes of missing data in such auxiliaries.

The applied example likely corresponds to outcome missingness mechanism 2 (an auxiliary used as a proxy for the missing outcome, which is thought to be MNAR) with auxiliary missingness mechanism 3 (MNAR) in our simulation study. Interpreted in the context of the simulation results, our applied example results suggest that the association between maternal smoking in pregnancy and offspring IQ at age 15 is biased towards the null by missing data, and that using an auxiliary with less missing data leads to a greater reduction in this bias. However, we caution against interpreting these results to mean that finding auxiliary variables with the least missing data should be an analyst’s only priority.

It is important to consider the nature of the relationship between an auxiliary variable and the variable being imputed (i.e., correctly specifying the imputation model)^3^. The applied example highlighted the importance of accounting for the non-linear relationship between KS4 score and IQ at age 15. Not accounting for this in the imputation model (as in models iii and iv) possibly led to overestimation of the effect.

The missing data mechanisms of the auxiliary variable and the variable to be imputed must also be considered in context of one another. In the applied example, the outcome and IQ age 8 were likely to be missing for similar groups of people, while the outcome and KS4 score were likely missing for two different groups. IQ (at age 8 and 15) was more likely to be missing for those with lower IQ (due to the socioeconomic patterning of participation in ALSPAC) whereas KS4 score was more likely to be missing for those attending an independent school (who were more likely to have higher IQ and KS4 scores). We cannot know if the difference between effect estimates obtained using imputation models including IQ at age 8 (model ii) versus KS4 score (including non-linear terms; model v) is due to using an auxiliary with more missing data or the auxiliary having a more similar missing data mechanism to the outcome (both occurred in model ii). This was not explored in the simulation study but warrants further investigation as overlap in missing data between variables may affect the ability of the auxiliary to predict missing values of the incomplete variable. At the extreme, if the auxiliary is either (1) always missing when Y is missing or (2) always missing when Y is observed, then the auxiliary will be no use whatsoever.

In the simulation study, missing data in the auxiliary and outcome were simulated under similar mechanisms; lower values of the outcome and auxiliary were both more likely to be missing. Therefore missing data in the outcome and auxiliary simultaneously is more likely in the simulation study and model ii of the applied example, but less likely in model v. Further, in the simulation study the proportion missing Z and Y simultaneously would have increased as the correlation between them increased. Despite this, in the simulation study bias was still reduced by the inclusion of the incomplete auxiliary in the imputation model but further investigation is needed to see if a greater reduction may have occurred with smaller overlap in missing data between the outcome and auxiliary.

The current study builds on prior work^10^ which explored the impact of 20% missing data in a MNAR auxiliary variable used as a proxy for an incomplete outcome (i.e., outcome mechanism 2), with missing data generated in a different way to our study. In the prior work missing data in the auxiliary did not have a substantial impact on effect estimates except when there was 80% missing data in the outcome. In the current study we explored greater proportions of missing data in the auxiliary and highlighted differences in bias reduction/introduction according to the missing data mechanism of the outcome and the auxiliary variable.

### Limitations

Our simulation scenarios were simple, consisting of only 3 to 4 continuous variables. The influence of missing auxiliary data on bias and efficiency in linear regression without non-linear terms may be different in models with interaction terms or logistic regression models because of non-collapsibility of the effect estimate.

We used a deterministic method to induce missing data in the outcome and auxiliary variables (i.e., an exact cut off for the CDF was used) as opposed to a stochastic process. The likely result is that the induced bias may be greater than would be observed in real world studies.

Missing data in analysis model variables was only assessed for an incomplete outcome, and not for incomplete exposure or confounding variables. Bias may be different when missing data occurs in such variables (compare results for β_*xy*_ to β_*yx*_ in the study by Collins et al.^7^), potentially as a result of Berkson error^24^.

We also used only a single mechanism for missing data in each variable in our simulations. More complex missing data mechanisms, including multiple reasons for missing data, may be more realistic. Bias in real world studies may therefore be greater or lesser than that found in the current study, dependent on the nature of the missing data mechanisms.

Finally, we only used a single auxiliary variable in our simulation study. In practice several auxiliary variables may be used, each of which may contain missing data. In this case the missing data for each auxiliary variable needs to be considered carefully. The exposure and auxiliaries may also not provide completely independent information, which would reduce the contribution of the auxiliary in predicting missing values.

## Conclusions

Careful consideration is required for the use of auxiliary variables in multiple imputation models. We suggest the following guidance:

- Use auxiliary variables that are completely observed, or almost completely observed.
- Explore the missing data mechanisms of incomplete auxiliary variables in addition to incomplete analysis model variables.
- Where auxiliary variables are incomplete, aim to use auxiliary variables that are MCAR or variables with evidence of being plausibly MAR, possibly conditional on another complete variable.

## Acknowledgements

This work was carried out using the computational facilities of the Advanced Computing Research Centre, University of Bristol - http://www.bris.ac.uk/acrc/. We are extremely grateful to all the families who took part in the ALSPAC study, the midwives for their help in recruiting them, and the whole ALSPAC team, which includes interviewers, computer and laboratory technicians, clerical workers, research scientists, volunteers, managers, receptionists, and nurses.

## Author contributions

PMD, JH and KT conceived the ideas and experimental design of the study. PMD performed the simulations, the applied analysis and wrote the initial and revised drafts of the manuscript. EC, RH, RC, JH and KT provided revisions to the scientific content and the stylistic/grammatical content of the manuscript.

## Data Availability

The ALSPAC data used in the applied example cannot be shared publicly for ethical reasons. The study website contains details of all available data through a fully searchable data dictionary (http://www.bristol.ac.uk/alspac/researchers/our-data/). The scripts and folder structure used to run the applied example analysis and simulation study can be found online at https://github.com/pmadleydowd/Missing_auxiliary_variables. Datasets for the simulation study are found within this repository.

## Additional Information

### Ethics approval

Ethical approval for the applied example (project B4170, searchable on https://proposals.epi.bristol.ac.uk/) was obtained from the ALSPAC Ethics and Law Committee and the Local Research Ethics Committees - http://www.bristol.ac.uk/alspac/researchers/research-ethics/.

### Consent to participate

Informed consent for the use of data collected via questionnaires and clinics was obtained from participants following the recommendations of the ALSPAC Ethics and Law Committee at the time. At age 18, study children were sent ‘fair processing’ materials describing ALSPAC’s intended use of education records and were given clear means to consent or object via a written form. Data were not extracted for participants who objected, or who were not sent fair processing materials.

## Abbreviations

ALSPAC: Avon longitudinal study of parents and children
CDF: cumulative distribution function
CRA: complete records analysis
DAG: directed acyclic graph
FCS: fully conditional specification
FMI: fraction of missing information
IQ: intelligence quotient
KS4: key stage 4
MAR: missing at random
MCAR: missing completely at random
MI: multiple imputation
MNAR: missing not at random

## References

1. Rubin DB. Inference and Missing Data. Biometrika. 1976;63(3):581–590. doi:DOI 10.1093/biomet/63.3.581

2. Rubin DB. Multiple imputation for nonresponse in surveys. New York: Wiley; 1987.

3. Curnow E, Carpenter JR, Heron JE, et al. Multiple imputation of missing data under missing at random: compatible imputation models are not sufficient to avoid bias if they are mis-specified. J Clin Epidemiol. Jun 19 2023;160:100–109. doi:10.1016/j.jclinepi.2023.06.011

4. Potthoff RF, Tudor GE, Pieper KS, Hasselblad V. Can one assess whether missing data are missing at random in medical studies? Stat Methods Med Res. Jun 2006;15(3):213–34. doi:10.1191/0962280206sm448oa

5. Schafer JL. Analysis of incomplete multivariate data. 1st ed. Chapman and Hall/CRC; 1997:444.

6. White IR, Royston P, Wood AM. Multiple imputation using chained equations: Issues and guidance for practice. Statistics in Medicine. Feb 20 2011;30(4):377–399. doi:10.1002/sim.4067

7. Collins LM, Schafer JL, Kam CM. A comparison of inclusive and restrictive strategies in modern missing data procedures. Psychol Methods. Dec 2001;6(4):330–51.

8. Hardt J, Herke M, Leonhart R. Auxiliary variables in multiple imputation in regression with missing X: a warning against including too many in small sample research. BMC Med Res Methodol. Dec 5 2012;12:184. doi:10.1186/1471-2288-12-184

9. Hughes RA, Heron J, Sterne JAC, Tilling K. Accounting for missing data in statistical analyses: multiple imputation is not always the answer. Int J Epidemiol. Mar 16 2019;doi:10.1093/ije/dyz032

10. Cornish RP, Macleod J, Carpenter JR, Tilling K. Multiple imputation using linked proxy outcome data resulted in important bias reduction and efficiency gains: a simulation study. Emerg Themes Epidemiol. 2017;14:14. doi:10.1186/s12982-017-0068-0

11. Thoemmes F, Rose N. A Cautious Note on Auxiliary Variables That Can Increase Bias in Missing Data Problems. Multivariate Behav Res. Sep-Oct 2014;49(5):443–59. doi:10.1080/00273171.2014.931799

12. Thoemmes F, Mohan K. Graphical Representation of Missing Data Problems. Structural Equation Modeling: A Multidisciplinary Journal. 2015/10/02 2015;22(4):631–642. doi:10.1080/10705511.2014.937378

13. Boyd A, Golding J, Macleod J, et al. Cohort Profile: the ‘children of the 90s’--the index offspring of the Avon Longitudinal Study of Parents and Children. International journal of epidemiology. 2013;42(1):111–27.

14. Fraser A, Macdonald-Wallis C, Tilling K, et al. Cohort Profile: the Avon Longitudinal Study of Parents and Children: ALSPAC mothers cohort. Int J Epidemiol. Feb 2013;42(1):97–110. doi:10.1093/ije/dys066

15. Major-Smith D, Heron J, Fraser A, Lawlor D, Golding J, Northstone K. The Avon Longitudinal Study of Parents and Children (ALSPAC): a 2022 update on the enrolled sample of mothers and the associated baseline data [version 1; peer review: awaiting peer review]. Wellcome Open Res. 2022;7(283)doi:10.12688/wellcomeopenres.18564.1

16. Donald HS, Giti C, Colin S. Concurrent Validity of the Wechsler Abbreviated Scale of Intelligence (WASI) with a Sample of Canadian Children. Canadian Journal of School Psychology. 2000/12/01 2000;16(1):87-94. doi:10.1177/082957350001600106

17. Wechsler D, Golombok S, Rust J. WISC-III UK. Sidcup, Kent: The Psychological Corporation. 1992;

18. Pearl J. Causality: models, reasoning, and inference. Cambridge University Press; 2000:xvi, 384 p.

19. Buuren Sv, Oudshoorn C. Multivariate imputation by chained equations: MICE V1. 0 user’s manual. 2000.

20. Cornish RP, Tilling K, Boyd A, Davies A, Macleod J. Using linked educational attainment data to reduce bias due to missing outcome data in estimates of the association between the duration of breastfeeding and IQ at 15 years. Int J Epidemiol. Jun 2015;44(3):937–45. doi:10.1093/ije/dyv035

21. Dempster AP, Laird NM, Rubin DB. Maximum likelihood from incomplete data via the EM algorithm. Journal of the royal statistical society Series B (methodological). 1977:1–38.

22. Madley-Dowd P, Hughes R, Tilling K, Heron J. The proportion of missing data should not be used to guide decisions on multiple imputation. J Clin Epidemiol. Jun 2019;110:63–73. doi:10.1016/j.jclinepi.2019.02.016

23. White IR. simsum: Analyses of simulation studies including Monte Carlo error. Stata Journal. 2010;10(3):369–385.

24. Haber G, Sampson J, Graubard B. Bias due to Berkson error: issues when using predicted values in place of observed covariates. Biostatistics. Oct 13 2021;22(4):858–872. doi:10.1093/biostatistics/kxaa002

