## Supplementary material for "Analyses using multiple imputation need to consider missing data in auxiliary variables"

### Supplementary materials

### Supplementary Tables

*Table S1: Descriptive statistics for ALSPAC sample meeting inclusion criteria in the applied example.*

| | Total N=11,780 | Non-smoker at 18 weeks' gestation N=8,812 | Current smoker at 18 weeks' gestation N=2,968 | p-value | Total with complete data in variable N=11,780 | Non-smoker at 18 weeks' gestation with complete data in variable N=8,812 | Current smoker at 18 weeks' gestation with complete data in variable N=2,968 |
|---|---|---|---|---|---|---|---|
| Offspring IQ at age 15 [a] | 94.6 (13.0) | 95.1 (13.0) | 92.0 (12.7) | <0.001 | 4,714 (40.0%) | 3,932 (44.6%) | 782 (26.3%) |
| Offspring IQ at age 8 [a] | 104.5 (16.4) | 105.2 (16.4) | 101.2 (16.0) | <0.001 | 6,567 (55.8%) | 5,328 (60.45) | 1,239 (41.7%) |
| Offspring KS4 score [a] | 325.2 (90.5) | 337.0 (84.1) | 289.4 (99.4) | <0.001 | 8,826 (74.9%) | 6,646 (75.4%) | 2,180 (75.5%) |
| Maternal age at birth [a] | 28.3 (4.9) | 28.9 (4.6) | 26.6 (5.1) | <0.001 | 11,780 (100%) | 8,812 (100%) | 2,968 (100%) |
| Parity Categories [b] | | | | <0.001 | 11,780 (100%) | 8,812 (100%) | 2,968 (100%) |
| 0 | 5,295 (44.9%) | 3,965 (45.0%) | 1,330 (44.8%) | | | | |
| 1 | 4,166 (35.4%) | 3,219 (36.5%) | 947 (31.9%) | | | | |
| 2+ | 2,319 (19.7%) | 1,628 (18.5%) | 691 (23.3%) | | | | |
| Maternal highest education [b] | | | | <0.001 | 11,780 (100%) | 8,812 (100%) | 2,968 (100%) |
| O-level or lower | 7567 (64.2%) | 5201 (59.0%) | 2366 (79.7%) | | | | |
| A-level/Degree or higher | 4,213 (35.8%) | 3,611 (41.0%) | 602 (20.3%) | | | | |
| Offspring assigned sex at birth [b] | | | | 0.011 | 11,780 (100%) | 8,812 (100%) | 2,968 (100%) |
| Female | 5,714 (48.5%) | 4,334 (49.2%) | 1,380 (46.5%) | | | | |
| Male | 6,066 (51.5%) | 4,478 (50.8%) | 1,588 (53.5%) | | | | |

[a] – mean (SD)

[b] – N(%)

[c] – T-tests applied to continuous variables and chi-squared tests applied to categorical variables

*Table S2: Average bias across simulations in complete records analysis and multiple imputation models excluding auxiliary variables.*

| Missing outcome mechanism | Missing auxiliary mechanism | Correlation between Z and Y | Bias (MCSE) in CRA model | Bias (MCSE) in MI model excluding auxiliaries |
|---|---|---|---|---|
| Mechanism 1 – MAR outcome given complete X and Z | Mechanism 1 – MCAR auxiliary | 0.1 | -0.047 (0.0008) | -0.046 (0.0008) |
| | | 0.3 | -0.139 (0.0008) | -0.139 (0.0008) |
| | | 0.5 | -0.231 (0.0008) | -0.231 (0.0008) |
| | | 0.7 | -0.323 (0.0008) | -0.323 (0.0008) |
| | Mechanism 2 – MAR auxiliary | 0.1 | -0.047 (0.0008) | -0.047 (0.0008) |
| | | 0.3 | -0.140 (0.0008) | -0.140 (0.0008) |
| | | 0.5 | -0.229 (0.0008) | -0.229 (0.0008) |
| | | 0.7 | -0.323 (0.0008) | -0.323 (0.0008) |
| | Mechanism 3 – MNAR auxiliary | 0.1 | -0.047 (0.0008) | -0.047 (0.0008) |
| | | 0.3 | -0.137 (0.0008) | -0.137 (0.0008) |
| | | 0.5 | -0.229 (0.0008) | -0.229 (0.0008) |
| | | 0.7 | -0.323 (0.0009) | -0.323 (0.0009) |
| Mechanism 2 – MNAR outcome | Mechanism 1 – MCAR auxiliary | 0.1 | -0.317 (0.0008) | -0.317 (0.0008) |
| | | 0.3 | -0.317 (0.0008) | -0.317 (0.0008) |
| | | 0.5 | -0.317 (0.0008) | -0.317 (0.0008) |
| | | 0.7 | -0.317 (0.0008) | -0.317 (0.0008) |
| | Mechanism 2 – MAR auxiliary | 0.1 | -0.318 (0.0009) | -0.318 (0.0009) |
| | | 0.3 | -0.318 (0.0008) | -0.318 (0.0008) |
| | | 0.5 | -0.316 (0.0008) | -0.316 (0.0008) |
| | | 0.7 | -0.317 (0.0008) | -0.317 (0.0008) |
| | Mechanism 3 – MNAR auxiliary | 0.1 | -0.317 (0.0008) | -0.317 (0.0008) |
| | | 0.3 | -0.317 (0.0008) | -0.317 (0.0008) |
| | | 0.5 | -0.316 (0.0008) | -0.316 (0.0008) |
| | | 0.7 | -0.318 (0.0008) | -0.318 (0.0008) |
| Mechanism 3 – MAR outcome given complete X | Mechanism 1 – MCAR auxiliary | 0.1 | -0.002 (0.0017) | -0.002 (0.0017) |
| | | 0.3 | -0.002 (0.0017) | -0.002 (0.0017) |
| | | 0.5 | -0.002 (0.0017) | -0.002 (0.0017) |
| | | 0.7 | -0.002 (0.0017) | -0.002 (0.0017) |
| | Mechanism 2 – MAR auxiliary | 0.1 | -0.003 (0.0017) | -0.003 (0.0017) |
| | | 0.3 | 0.001 (0.0017) | 0.001 (0.0017) |
| | | 0.5 | -0.001 (0.0017) | -0.001 (0.0017) |
| | | 0.7 | -0.002 (0.0017) | -0.002 (0.0017) |
| | Mechanism 3 – MNAR auxiliary | 0.1 | 0.001 (0.0017) | 0.001 (0.0017) |
| | | 0.3 | -0.001 (0.0017) | -0.001 (0.0017) |
| | | 0.5 | 0.003 (0.0016) | 0.003 (0.0017) |
| | | 0.7 | -0.002 (0.0017) | -0.003 (0.0017) |

CRA = Complete Records Analysis; MAR = Missing at random; MCAR = Missing completely at random; MCSE = Monte Carlo Standard Error; MI = Multiple imputation; MNAR = Missing not at random

#### Assessment of the assumed relationship between variables and missing data in the applied example

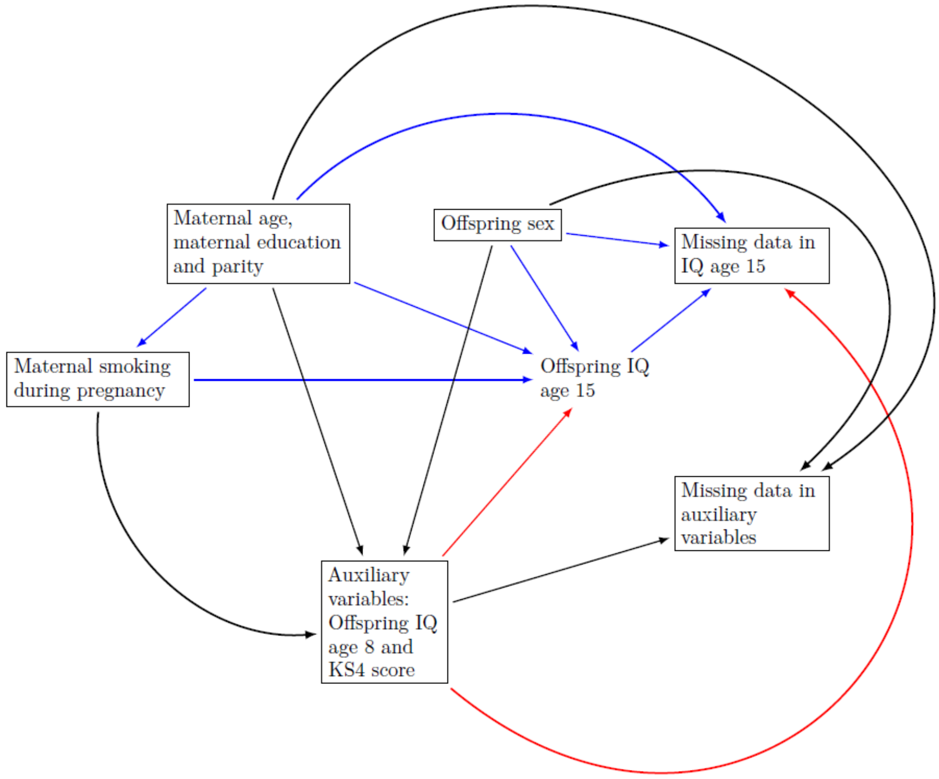

*Figure S1: Directed acyclic graph for the applied example. Lines in blue show relationships between variables that are part of the complete records analysis model. Lines in red represent a biasing pathway that is closed through the inclusion of the auxiliary variables in the imputation model.*

Figure S1 shows a DAG of the assumed relationship between variables and missing data indicators in the applied example. The DAG has been simplified, which suffices for our purposes, and not all relationships are represented. Below we justify the assumed relationship between each node and detail some of the assumed parametric relationships that cannot be captured by DAGs (because DAGs are not parametric in nature).

##### Confounding

Confounding variables were recorded using maternal questionnaire during the pregnancy period. Maternal age, parity and education proxy for socioeconomic position which is associated with maternal smoking in pregnancy [1,2] and offspring IQ [3-5]. Offspring sex may explain some of the variation in IQ [6], likely as a result of sex-differences in conscientiousness or school children's ability to take cognitive tests rather than sex differences in average intelligence, though sex will not influence the exposure. As such we do not consider this a confounder. It can, however, be used to reduce standard error of the exposure coefficient.

#### Auxiliary variables

Figure S1 shows that the auxiliary variable offspring IQ at age 8 is on the causal pathway between the exposure, maternal smoking during pregnancy, and the outcome, IQ at age 15. Inclusion of IQ at age 8 in the analysis model would therefore lead to estimation of a direct effect of exposure on outcome not via IQ at age 8. Thus we do not include IQ at age 8 in the analysis model, as we wish to estimate the total effect of exposure on outcome.

In contrast, KS4 scores are obtained after the measurement of the outcome at the age of 16. This is not represented by the DAG as a result of simplification. Readers should note that when considering KS4 score as the auxiliary the arrow between offspring IQ at age 15 and the auxiliary variable is the reverse to that presented in Figure S1 (i.e., replace IQ at age 15 ← IQ at age 8 with IQ at age 15 → KS4 score). Adjustment for KS4 score in the analysis model would result in collider bias as the variable is a descendent of both the exposure and the outcome.

The auxiliary variables therefore have to act as proxies for missing outcome information within the imputation model [7,8] to ensure estimation of the effect we are interested in and to prevent collider bias. Inclusion of either IQ at age 8 or KS4 score in the imputation model will remove a portion of the relationship between the outcome and its own missing data indicator thereby reducing bias from missing data (as a result of blocking the open path in red along IQ at age 15 ← IQ at age 8 → missing data in IQ at age 15, or along a path that is not included in the simplified DAG that may look something like: IQ at age 15 ← underlying academic ability proxied for by KS4 scores at age 16 → missing data in IQ at age 15). It is important to note that inclusion of these auxiliary variables in the imputation model does not completely sever the relationship between the outcome and its missing data indicator as a result of the direct arrow from IQ at age 15 to missing data in IQ at age 15 (i.e., IQ at age 15 is MNAR). Some bias from missing data will therefore remain and is unable to be removed without additional data or additional assumptions.

In Figure S1 we did not include an arrow from IQ at age 15 to missing data in auxiliary variables as IQ at age 15 cannot cause missing data in IQ captured at age 8. We do note however that IQ at age 15 could plausibly cause missing KS4 scores, captured at age 16, and that an arrow from IQ at age 15 to the auxiliary variable node has been omitted from the DAG. Combined with the fact that we consider it likely that both auxiliary variables are MNAR, as they may be a cause of their own missingness, it is possible that including these auxiliary variables in the imputation model may create pathways from the outcome to the missing data indicator for the auxiliary variables, thereby introducing some bias.

More pathways would exist for KS4 score as an auxiliary than IQ at age 8, if IQ at age 15 were to cause missing data in KS4 score, though any bias may be smaller due to the lower proportion of missing data in KS4 score than IQ at age 8. We consider the amount of bias introduced by inclusion of the auxiliary variables to be less than the amount removed by closing pathways from IQ at age 15 to its own missing data indicator.

It has previously been shown that IQ and KS4 attainment are related in a non-linear way and that the relationship varies according to maternal education [8] – this cannot be represented within the DAG but needs to be considered when imputing values of IQ from KS4 attainment.

#### Missing data mechanisms

We attempt to provide evidence for the assumed relationships presented in Figure S1 by providing ORs in Table S3 for missing data in the outcome and each of the auxiliary variables according to each variable included in the applied example. It is important to note that the relationships between variables with missing data and the probability of missing data cannot be verified (and further that we have excluded participants with missing confounder data for simplicity of the example and so these results may not hold for ALSPAC more generally).

Table S3 shows that missing data in IQ variables (at age 8 and 15) was more likely among offspring with lower IQ and lower KS4 scores. Missing IQ was also more likely for offspring of mothers who were younger, had higher parity and had lower education levels. In contrast, KS4 scores were more likely to be missing among offspring with higher IQ at age 15 (but not age 8), and had mothers with higher levels of education. Parity and maternal age did not have strong evidence for an association with missing data in KS4 scores. Male sex and having a mother who smoked during pregnancy was associated with increased odds of missing data in IQ variables (at both ages, though the effect of sex was stronger at age 15 than age 8) and KS4 score. Maternal smoking during pregnancy was a strong predictor of missing data in IQ at age 15 (OR = 1.79; 95% CI = 1.62-1.96), missing data in IQ at age 8 (OR = 1.64; 95% CI = 1.50-1.79) and missing data in KS4 score (OR = 1.22; 95% CI = 1.10-1.34) after adjusting for maternal socioeconomic variables (education level, age and parity) suggesting that there may be a direct link from the exposure variable to missing data in the outcome and missing data in the auxiliary variables that cannot be accounted for by our measured confounding variables.

Prior work in ALSPAC has shown that socioeconomic position and sex predicted offspring participation in ALSPAC in late teenage years [9]. The outcome (IQ at age 15) may therefore be more likely to be missing for those with lower IQ due to the relationship between lower socioeconomic position and lower IQ scores [3-5]. In contrast linked educational data (capturing KS4 scores) could not be obtained for participants attending an independent or private school. These are schools where students are from a higher socioeconomic position, are more likely to have higher educational attainment and therefore have higher KS4 scores. The relationship between offspring sex and missing data in IQ at age 8 was smaller in magnitude than the relationship with missing data at age 15. This may be because participants are more likely to be brought to clinics for IQ testing by their parents. Missing data in KS4 scores was more likely among offspring who's assigned sex at birth was male. This may be due to a larger proportion of males than females permanently excluded from school [10].

*Table S3: OR for missing data in the outcome and auxiliary variables according to the variables included in the applied example.*

| | OR (95% CI) for missing data in IQ at age 15 | OR (95% CI) for missing data in IQ at age 8 | OR (95% CI) for missing data in KS4 score |
|---|---|---|---|
| Offspring IQ at age 15 | - | 0.97 (0.96-0.98) | 1.02 (1.01-1.03) |
| Offspring IQ at age 8 | 0.98 (0.97-0.98) | - | 1.00 (1.00-1.01) |
| Offspring KS4 score | 0.99 (0.99-0.99) | 0.99 (0.99-0.99) | - |
| Maternal smoking during pregnancy | 2.35 (2.15-2.57) | 2.23 (2.06-2.41) | 1.13 (1.04-1.24) |
| Maternal age at birth | 0.92 (0.92-0.93) | 0.91 (0.90-0.91) | 1.01 (1.00-1.01) |
| Parity Categories | | | |
| 0 | Ref | Ref | Ref |
| 1 | 1.19 (1.10-1.29) | 1.05 (0.97-1.14) | 0.85 (0.87-1.04) |
| 2+ | 1.65 (1.50-1.83) | 1.43 (1.30-1.57) | 1.04 (0.94-1.15) |
| Maternal highest education | | | |
| O-level or lower | Ref | Ref | Ref |
| A-level/Degree or higher | 0.45 (0.42-0.49) | 0.44 (0.41-0.48) | 1.37 (1.26-1.49) |
| Offspring assigned sex at birth | | | |
| Female | Ref | Ref | Ref |
| Male | 1.26 (1.18-1.35) | 1.12 (1.05-1.20) | 1.28 (1.19-1.37) |

### Fraction of missing information (FMI) results

#### Applied example

The exposure coefficient had the largest FMI of all covariates in all MI models. Inclusion of auxiliary variables led to a reduction in the exposure FMI compared to MI excluding auxiliary variables. This shows that, despite the missing data in the auxiliary variables, their inclusion provided more information about the relationship between the exposure and missing outcome variable than excluding them from the imputation model. Inclusion of non-linear terms in the imputation model when KS4 score was used as an auxiliary led to an increase in the FMI (compare model iii to model v) suggesting that correct specification of the model does not necessarily reduce the FMI.

#### Simulation study

We present the FMI for the exposure coefficient for all outcome and auxiliary missing data mechanism combinations in Figure S2. The FMI of MI models excluding auxiliary variables (horizontal dashed line in each plot) was largest for data simulated under outcome missingness mechanism 3 and smallest under mechanism 1. For all missing outcome mechanisms, lower correlation between Z and Y (reflecting a weaker ability of Z to predict Y) led to higher FMI in MI models including auxiliary variables with no missing auxiliary data. Under outcome mechanism 1 (Figure S2 A-C) inclusion of complete auxiliaries with low correlation between Z and Y (0.3 or 0.1) led to FMI values that were greater than MI models excluding auxiliary variables. When auxiliary variables were MCAR or MNAR, as the proportion of missing data in the auxiliary increased, the FMI of models with different correlations between Z and Y all converged to the value obtained in models excluding auxiliary variables. For MAR auxiliary data, the FMI of all MI models including auxiliary variables converged to a value greater than that observed in MI models excluding auxiliary variables. The behaviour of the FMI with increasing proportion of missing auxiliary data was similar between outcome mechanism 2 (Figure S2 D-F) and outcome mechanism 3 (Figure S2 G-I). In both cases, for all missing auxiliary mechanisms, low correlation between Z and Y in MI including auxiliary variables led to FMI values closer to the FMI of MI excluding auxiliary variables at all proportions of missing data. As the proportion of missing data in the auxiliary variable increased the FMI of models including auxiliary variables converged to the value of MI models excluding auxiliary variables.

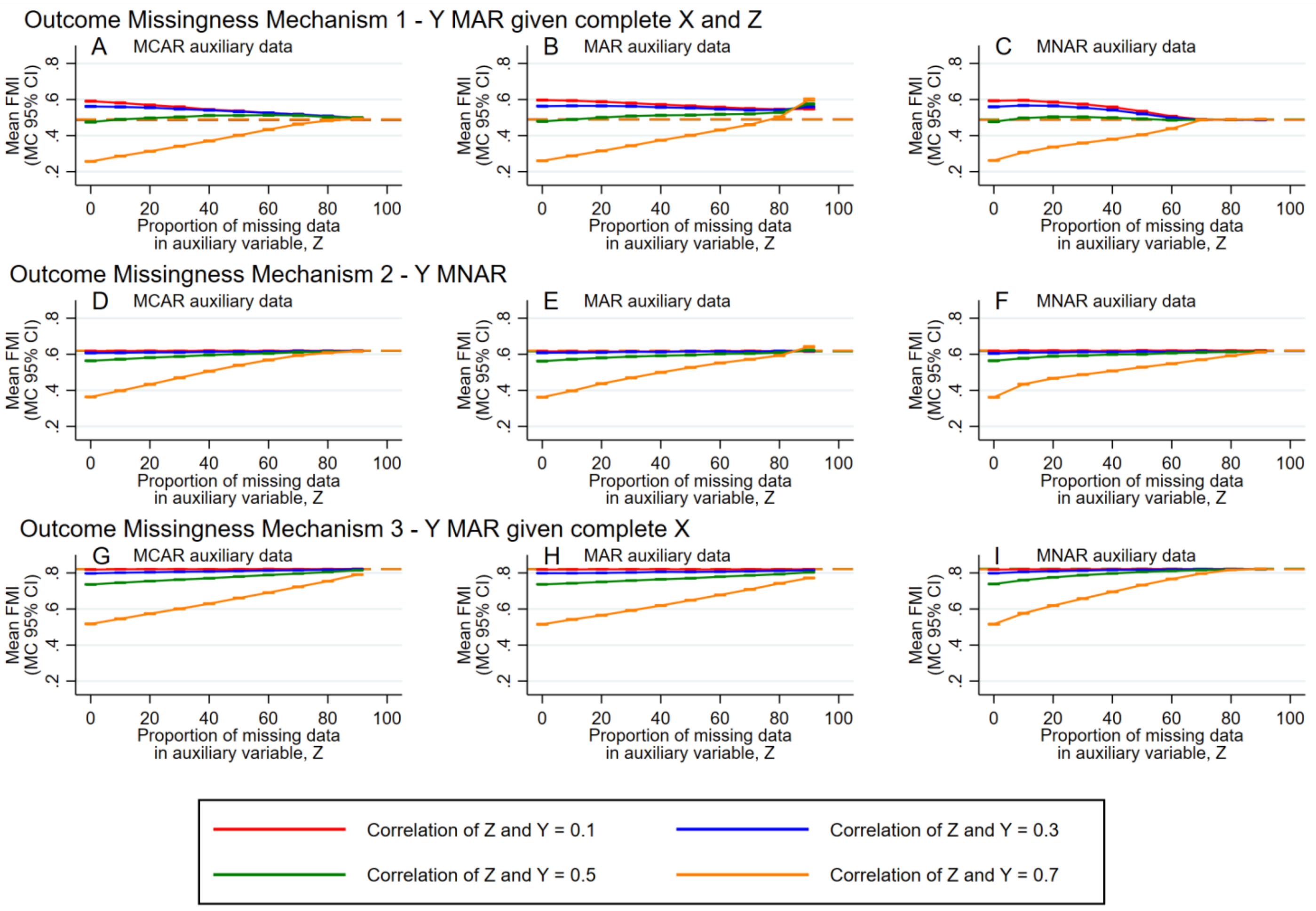

*Figure S2: Plots of mean FMI of the effect estimate of the exposure, X, across simulations against the proportion of missing data in the auxiliary variable, Z, for each level of correlation between the auxiliary, and outcome, Y. Solid lines correspond to FMI of MI including auxiliary variables while dashed lines correspond to FMI of MI excluding auxiliary variables. FMI = fraction of missing information, CI = confidence interval, MC = Monte Carlo, MCAR = missing completely at random, MAR = missing at random, MNAR = missing not at random*

### Applied example sensitivity analyses

#### Distribution of missing data according to included vs. excluded participants

Supplementary Table S4 shows descriptives for included versus excluded participants with missing values in the confounders. Missing data in the outcome and auxiliary variables was substantially more likely when confounder variables were also missing.

*Table S4: Distribution of observed and missing data in the sample with all confounders observed vs the sample with some missing confounder information. Participants with missing values in the confounders were more likely to have a larger number of missing variables for the outcome, exposure and auxiliary variables.*

| | Total<br>N=13,826 | All confounders observed (included)<br>N=11,780 | Some confounders missing (excluded)<br>N=2,046 |
|---|---|---|---|
| Offspring IQ at age 15 | | | |
| Observed | 4,983 (36.0%) | 4,714 (40.0%) | 269 (13.1%) |
| Missing | 8,843 (64.0%) | 7,066 (60.0%) | 1,777 (86.9%) |
| Offspring IQ at age 8 | | | |
| Observed | 6,986 (50.5%) | 6,567 (55.7%) | 419 (20.5%) |
| Missing | 6,840 (49.5%) | 5,213 (44.3%) | 1,627 (79.5%) |
| Offspring KS4 score | | | |
| Observed | 10,139 (73.3%) | 8,826 (74.9%) | 1,313 (64.2%) |
| Missing | 3,687 (26.7%) | 2,954 (25.1%) | 733 (35.8%) |
| Maternal smoking during pregnancy | | | |
| Observed | 13,010 (94.1%) | 11,780 (100.0%) | 1,230 (60.1%) |
| Missing | 816 (5.9%) | 0 (0.0%) | 816 (39.9%) |
| Maternal age | | | |
| Observed | 13,826 (100.0%) | 11,780 (100.0%) | 2,046 (100.0%) |
| Missing | 0 (0.0%) | 0 (0.0%) | 0 (0.0%) |
| Parity | | | |
| Observed | 12,784 (92.5%) | 11,780 (100.0%) | 1,004 (49.1%) |
| Missing | 1,042 (7.5%) | 0 (0.0%) | 1,042 (50.9%) |
| Maternal education | | | |
| Observed | 12,276 (88.8%) | 11,780 (100.0%) | 496 (24.2%) |
| Missing | 1,550 (11.2%) | 0 (0.0%) | 1,550 (75.8%) |

#### Reducing the number of imputations used in the applied example

The number of imputations used in the applied example was larger than would feasibly be used in an applied study. We therefore reran analyses using 100 imputations. The results, which do not differ substantially from models using 1000 imputations, are presented in Supplementary Table S5.

*Table S5: Model results for applied MI analyses using 100 imputations.*

| Model | Exposure coefficient (s.e.) [a] | 95% CI | FMI for exposure coefficient | Largest FMI of all covariates |
|---|---|---|---|---|
| CRA | -0.87 (0.485) | -1.82, 0.08 | - | - |
| i) MI excluding auxiliaries | -0.81 (0.483) | -1.76, 0.15 | 0.70 | 0.70 |
| ii) MI with IQ at age 8 as an auxiliary | -1.13 (0.448) | -2.01,-0.24 | 0.65 | 0.65 |
| iii) MI with KS4 score as an auxiliary | -1.99 (0.431) | -2.83,-1.14 | 0.57 | 0.57 |
| iv) MI with IQ at age 8 and KS4 score as auxiliaries | -1.94 (0.428) | -2.78,-1.09 | 0.58 | 0.58 |
| v) MI with KS4 score cubed as an auxiliary (including multiplicative term [b]) | -1.52 (0.467) | -2.44,-0.60 | 0.67 | 0.67 |
| vi) MI with IQ at age 8 and KS4 score cubed as auxiliaries (including multiplicative term [b]) | -1.45 (0.407) | -2.25,-0.65 | 0.57 | 0.57 |

[a] Mean difference in IQ at age 15 among offspring of maternal smokers during pregnancy compared to non-smokers during pregnancy

[b] Multiplicative term between KS4 score cubed and maternal education

### Simulation study sensitivity analyses

#### Methods

In sensitivity analyses we explored the impact of i) the proportion of missing data in the outcome and ii) the strength of association between X and Y relative to Z and Y. We therefore repeated the simulation study using i) 75% missing data in the outcome and ii) a correlation between X and Y of 0.2. Our sensitivity analyses were conducted under MAR auxiliary data only (missing auxiliary mechanism 2) for missing outcome mechanisms 1 and 2, and under MNAR auxiliary data only (missing auxiliary mechanism 3) for missing outcome mechanism 3. The mechanisms explored were selected based on the results of the main simulation results as the most important scenarios to understand. For mechanism 1 and 2 we chose to assess sensitivity under MAR auxiliary data as these mechanisms are biased under a CRA and we want to understand how the change in parameters influenced our ability to reduce bias. For mechanism 3 there was no bias in CRA but there was under some MI models, and so we wanted to understand how modification of the parameters influenced the quantity of bias introduced.

We additionally explored the impact of excluding W from the imputation model for Z under missing auxiliary mechanism 2 for all missing outcome mechanisms. This scenario is another example of a MNAR auxiliary missing data mechanism as Z was MAR conditional on W.

#### Results

##### Bias for sensitivity analysis i) and ii)

The bias under CRA and MI without auxiliaries for sensitivity analyses assessing the impact of i) the proportion of missing data in the outcome and ii) the strength of association between X and Y relative to Z and Y are presented in Table S6. Under outcome missingness mechanism 1 and 2, bias in CRA (and equivalently bias in MI models excluding auxiliary variables) was increased when the proportion of missing data in the outcome variable was increased from 50% to 75%. In contrast, when the correlation between X and Y was reduced from 0.6 to 0.2, the bias in CRA remained approximately equivalent under missing outcome mechanism 1 but was reduced under missing outcome mechanism 2. Bias was approximately equal to zero for all CRA models under missing outcome mechanism 3.

Figure S3 presents plots of bias for MI models in sensitivity analyses i) and ii). Under outcome mechanism 1 (Y MAR conditional on complete Z and X), bias relative to CRA remained approximately the same as in the main analyses when the proportion of missing outcome was increased (compare

Figure S3A to Figure 2B) and when the correlation between X and Y was reduced (compare Figure S3D to Figure 2B), though the reduction in relative bias for the largest correlation between Z and Y was less pronounced in sensitivity analysis ii). This suggests that MI is consistent in its ability to reduce bias when both the outcome and auxiliary variables are MAR, irrespective of changes in the proportion of missing outcome data or reductions in the strength of association between exposure and outcome.

For outcome mechanism 2 (Y MNAR), bias relative to CRA increased when the proportion of missing outcome data was increased from 50 to 75% (compare Figure S3B to Figure 2E) and when the correlation between X and Y was reduced from 0.6 to 0.2 (compare Figure S3E to Figure 2E). This was true for all proportions of missing auxiliary data and all strengths of correlation between Y and Z. This suggests that when we use auxiliary variables to act as proxy variables for the outcome, the ability of MI to reduce bias relative to CRA is influenced by other factors, even when the auxiliary is MAR.

For outcome mechanism 3 (Y MAR conditional on complete X), where there was zero bias in the CRA, the absolute bias in sensitivity analysis for MI using MNAR auxiliary variables was smaller relative to the results of the primary analyses. This was true when the proportion of missing outcome data increased (compare Figure S3C to Figure 2I) and when the correlation between X and Y was reduced (compare Figure S3F to Figure 2I). The maximum quantity of bias remained at around 50% missing data in the auxiliary variable, even when the proportion of missing data in the outcome was increased to be greater than 50%, suggesting that this is a pattern not introduced as a result of our simulation design choice to use 50% missing outcome data.

#### FMI for sensitivity analyses i) and ii)

Plots of the FMI for sensitivity analyses i) and ii) are presented in Figure S4. Under missing outcome mechanism 1 (MAR outcome), in primary and sensitivity analyses MI models with large proportions of MAR auxiliary data lead to FMI values that were larger than values from models in which auxiliary variables were excluded. The proportion of missing auxiliary data at which including the auxiliary variable made the FMI worse than if the auxiliary was excluded, varied according to factors such as how much missing data in the outcome there was (compare Figure S4A to Figure S2B) and the strength of association between exposure and outcome (compare Figure S4D to Figure S2B). The FMI increased for all correlations between Z and Y and all proportions of missing auxiliary variable when the proportion of the outcome that was missing increased from 50% to 75%, highlighting the impact of the proportion of missing data in the analysis model on the FMI, irrespective of the amount of missing data in the auxiliary variable. When the correlation of X and Y was reduced to 0.2, the reduction in FMI at low proportions of missing auxiliary data with higher correlations between Z and

Y was smaller, suggesting that there is a reduced ability for the auxiliary to retain missing information on the exposure-outcome effect estimate when this effect is truly smaller.

When the outcome was MNAR (missing outcome mechanism 2) in primary analyses the FMI was approximately equal to the MI model excluding auxiliary variables for weaker correlations (≤0.3) between auxiliary and outcome (see Figure S2E). For stronger correlations (≥0.5) the FMI was lower than the MI model excluding auxiliary variables while the proportion of missing data in the auxiliary variable remained lower than 50%. Sensitivity analysis showed that a similar pattern was found when the proportion of missing data in the outcome increased to 75% (compare Figure S4B to Figure S2E) or when the correlation between exposure and outcome was reduced (compare Figure S4E to Figure S2E).

Under mechanism 3 where outcome data were MAR conditional on complete exposure information, the FMI behaved in a comparable manner to mechanism 2, though the maximum value of the FMI was greater than that observed under mechanism 1 or 2 (compare Plot C to A and B and Plot F to D and E in Figure S4). The FMI was greater in sensitivity analyses where the proportion of missing outcome was increased from 50% to 75% (compare Figure S4C to Figure S2I) but remained at a similar value to primary analyses when the correlation between X and Y was reduced (compare Figure S4F to Figure S2I).

#### Bias and FMI when excluding W from the imputation model under missing auxiliary mechanism 2

We additionally assessed the impact of excluding W from the imputation model under missing auxiliary mechanism 2 in which Z was MAR conditional on W. Plots of bias for this sensitivity analysis are presented in Figure S5 and plots of the FMI are presented in Figure S6. When W was excluded from the imputation model bias was increased at all proportions of missing data for all missing outcome mechanisms, compared to when W was included in the imputation model (thereby making Z MAR). Inclusion of W in the imputation model led to FMI values that were larger than in MI models excluding all auxiliaries for large proportions of missing data in Z (>80%) under missing outcome mechanism 1 and 2; this was more pronounced when there were stronger correlations between Z and Y. Exclusion of W from the imputation model lead to the FMI converging to the FMI value of models excluding all auxiliaries as the proportion of missing data in Z increased.

*Table S6: Sensitivity analysis – average bias across simulations in complete records analysis and MI models excluding auxiliary variables.*

| Missing auxiliary mechanism | Missing outcome mechanism | Sensitivity analysis | Correlation between Z and Y | Bias (MCSE) in CRA model | Bias (MCSE) in MI model excluding auxiliaries |
|---|---|---|---|---|---|
| Mechanism 2 – MAR auxiliary given complete W | Mechanism 1 – MAR outcome given complete X and Z | 1 – more missing outcome (75%) | 0.1 | -0.060 (0.0011) | -0.060 (0.0011) |
| | | | 0.3 | -0.177 (0.0012) | -0.177 (0.0012) |
| | | | 0.5 | -0.295 (0.0013) | -0.295 (0.0013) |
| | | | 0.7 | -0.414 (0.0012) | -0.414 (0.0012) |
| | | 2 – smaller X-Y correlation (0.2) | 0.1 | -0.046 (0.0010) | -0.046 (0.0010) |
| | | | 0.3 | -0.138 (0.0010) | -0.138 (0.0010) |
| | | | 0.5 | -0.230 (0.0010) | -0.230 (0.0010) |
| | | | 0.7 | -0.321 (0.0010) | -0.321 (0.0010) |
| | Mechanism 2 – MNAR outcome | 1 – more missing outcome (75%) | 0.1 | -0.400 (0.0012) | -0.400 (0.0012) |
| | | | 0.3 | -0.399 (0.0012) | -0.398 (0.0012) |
| | | | 0.5 | -0.402 (0.0011) | -0.402 (0.0011) |
| | | | 0.7 | -0.401 (0.0011) | -0.401 (0.0011) |
| | | 2 – smaller X-Y correlation (0.2) | 0.1 | -0.126 (0.0010) | -0.126 (0.0010) |
| | | | 0.3 | -0.126 (0.0010) | -0.126 (0.0010) |
| | | | 0.5 | -0.127 (0.0009) | -0.127 (0.0010) |
| | | | 0.7 | -0.126 (0.0010) | -0.126 (0.0010) |
| Mechanism 3 – MNAR auxiliary | Mechanism 3 – MAR outcome given complete X | 1 – more missing outcome (75%) | 0.1 | 0.005 (0.0032) | 0.004 (0.0032) |
| | | | 0.3 | 0.004 (0.0032) | 0.004 (0.0033) |
| | | | 0.5 | 0.003 (0.0032) | 0.002 (0.0032) |
| | | | 0.7 | 0.001 (0.0032) | 0.001 (0.0032) |
| | | 2 – smaller X-Y correlation (0.2) | 0.1 | -0.000 (0.0022) | 0.000 (0.0022) |
| | | | 0.3 | 0.003 (0.0021) | 0.003 (0.0021) |
| | | | 0.5 | -0.000 (0.0021) | -0.000 (0.0021) |
| | | | 0.7 | -0.002 (0.0021) | -0.002 (0.0021) |

CRA = Complete Records Analysis; MAR = Missing at random; MCAR = Missing completely at random; MCSE = Monte Carlo Standard Error; MI = Multiple imputation; MNAR = Missing not at random

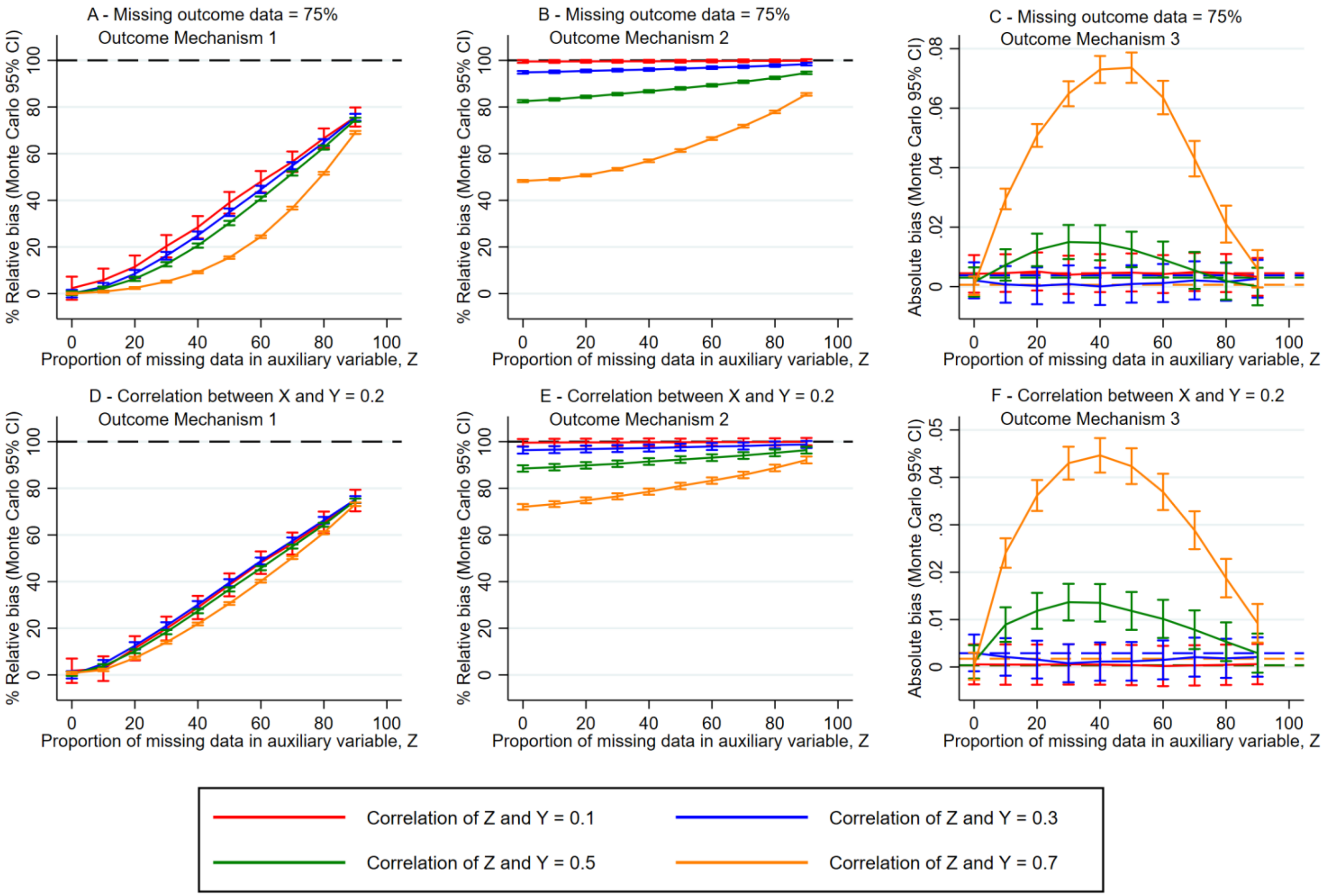

*Figure S3: Plots of bias (relative to CCA for outcome mechanisms 1 and 2, and absolute for outcome mechanism 3) for the exposure (X) coefficient against the proportion of missing data in Z for sensitivity analysis that i) increased the missing data in the outcome to 75% (plots A-C) and ii) decreased the correlation between X and Y to 0.2 (plots D-E). The missing auxiliary mechanism is mechanism 2 for plots A, B, D and E and mechanism 3 for plots C and F.*

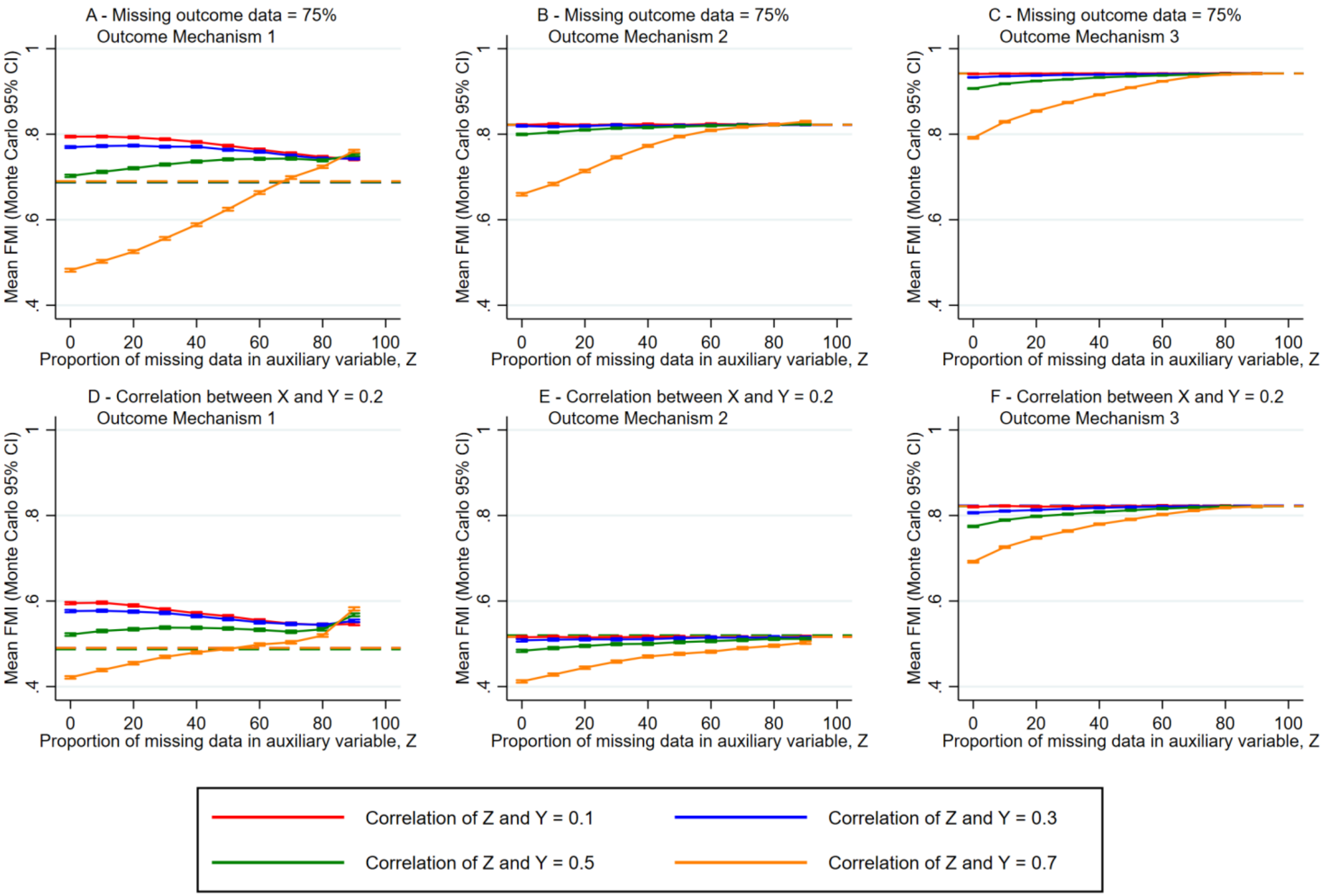

*Figure S4: Plots of the FMI for the exposure (X) coefficient against the proportion of missing data in Z for sensitivity analysis that i) increased the missing data in the outcome to 75% (plots A-C) and ii) decreased the correlation between X and Y to 0.2 (plots D-E). The missing auxiliary mechanism is mechanism 2 for plots A, B, D and E and mechanism 3 for plots C and F.*

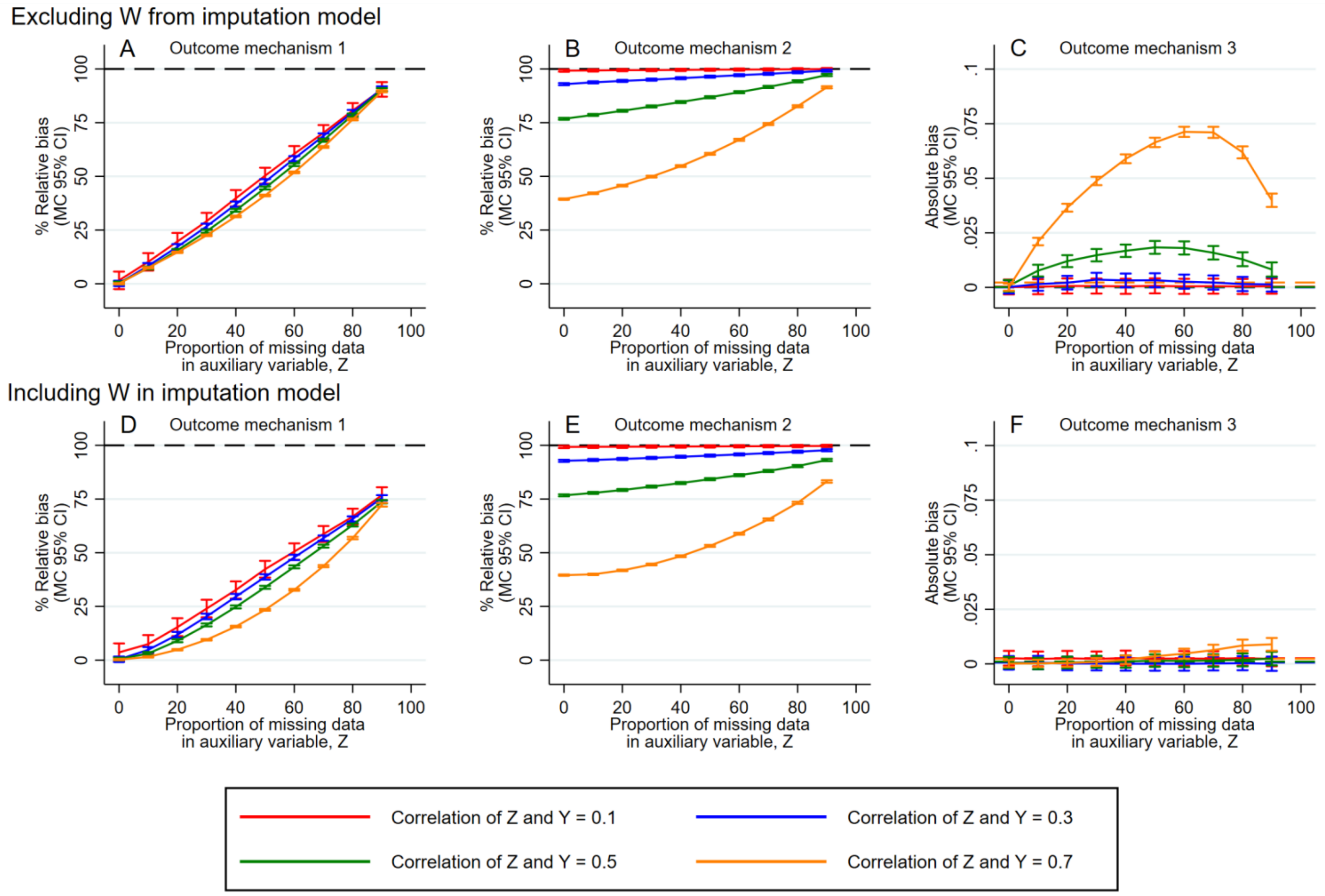

*Figure S5: Plots of bias (relative to CCA for outcome mechanisms 1 and 2, and absolute for outcome mechanism 3) for the exposure (X) coefficient against the proportion of missing data in Z for sensitivity analysis that excluded W from the imputation model under missing auxiliary mechanism 2 (plots A-C). For ease of comparison, we show the results of models in which W was included in the imputation model for missing auxiliary mechanism (plots D-F; these are also shown as plots B,E and H in Figure 2 of the main text).*

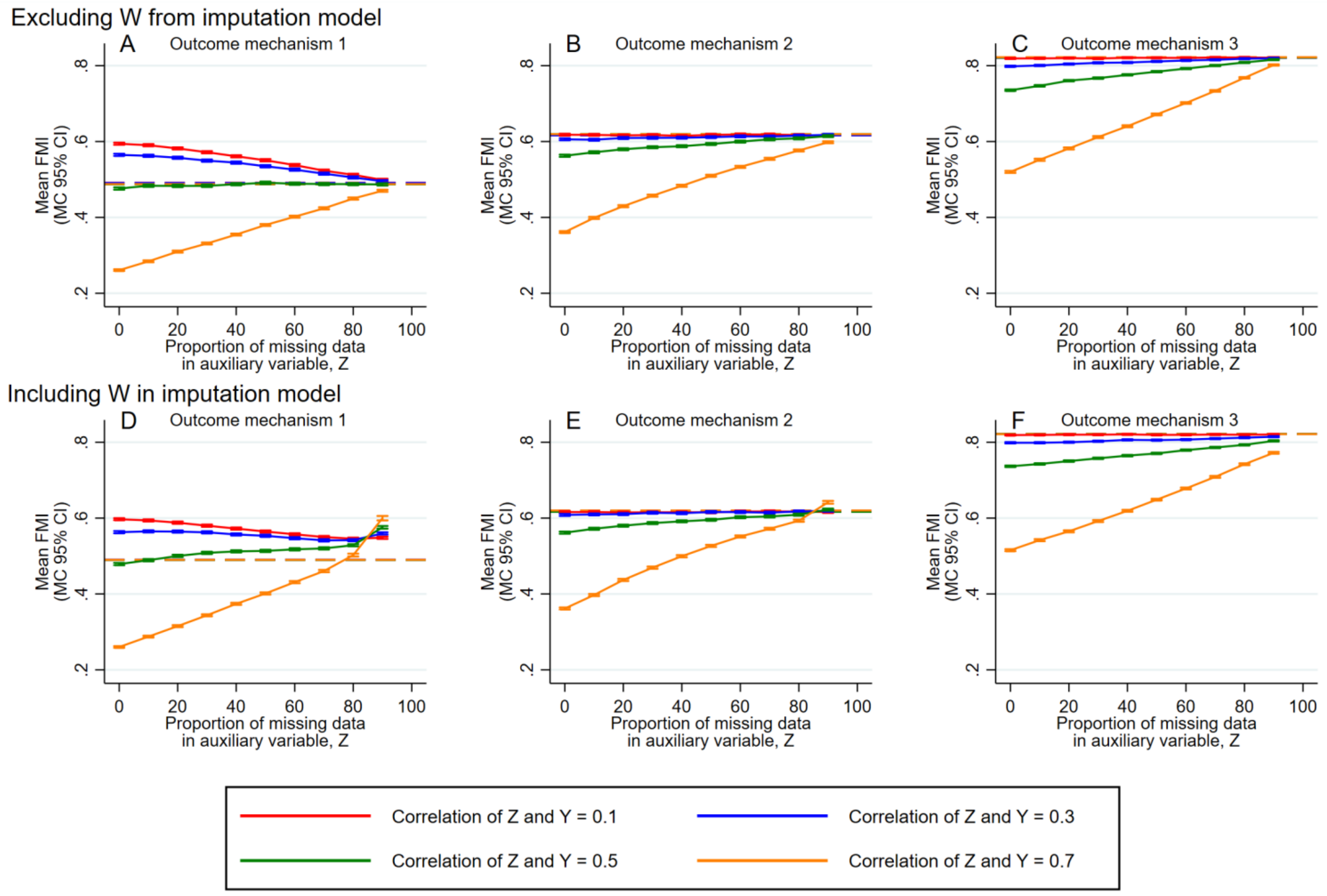

*Figure S6: Plots of the FMI for the exposure (X) coefficient against the proportion of missing data in Z for sensitivity analysis that excluded W from the imputation model under missing auxiliary mechanism 2 (plots A-C). For ease of comparison, we show the results of models in which W was included in the imputation model for missing auxiliary mechanism (plots D-F; these are also shown as plots B,E and H in Figure S3).*

### References

1. Hiscock R, Bauld L, Amos A, Fidler JA, Munafo M. Socioeconomic status and smoking: a review. *Ann N Y Acad Sci.* 2012;1248:107-123.
2. Lu Y, Tong S, Oldenburg B. Determinants of smoking and cessation during and after pregnancy. *Health Promotional International.* 2001;16(4):355-365.
3. Hair NL, Hanson JL, Wolfe BL, Pollak SD. Association of Child Poverty, Brain Development, and Academic Achievement. *JAMA Pediatrics.* 2015;169(9):822-829.
4. Eriksen HL, Kesmodel US, Underbjerg M, Kilburn TR, Bertrand J, Mortensen EL. Predictors of intelligence at the age of 5: family, pregnancy and birth characteristics, postnatal influences, and postnatal growth. *PLoS One.* 2013;8(11):e79200.
5. Hackman DA, Gallop R, Evans GW, Farah MJ. Socioeconomic status and executive function: developmental trajectories and mediation. *Dev Sci.* 2015;18(5):686-702.
6. Lynn R, Kanazawa S. A longitudinal study of sex differences in intelligence at ages 7, 11 and 16 years. *Pers Indiv Differ.* 2011;51(3):321-324.
7. Cornish RP, Macleod J, Carpenter JR, Tilling K. Multiple imputation using linked proxy outcome data resulted in important bias reduction and efficiency gains: a simulation study. *Emerg Themes Epidemiol.* 2017;14:14.
8. Cornish RP, Tilling K, Boyd A, Davies A, Macleod J. Using linked educational attainment data to reduce bias due to missing outcome data in estimates of the association between the duration of breastfeeding and IQ at 15 years. *Int J Epidemiol.* 2015;44(3):937-945.
9. Boyd A, Golding J, Macleod J, et al. Cohort Profile: the 'children of the 90s'--the index offspring of the Avon Longitudinal Study of Parents and Children. *International journal of epidemiology.* 2013;42(1):111-127.
10. Department for Education. Permanent and fixed-period exclusions in England: 2008 to 2009. School discipline and exclusions Web site. https://www.gov.uk/government/statistics/permanent-and-fixed-period-exclusions-in-england-academic-year-2008-to-2009. Published 2010. Accessed 31 August, 2023.
